# Differential age-specific effects of LDL cholesterol and body mass index on the risk of coronary heart disease

**DOI:** 10.1101/2023.02.21.23286273

**Authors:** Jun Xiao, Liangwan Chen, Weimin Ye, Wuqing Huang

**Affiliations:** Department of Cardiovascular Surgery, Fujian Medical University Union Hospital, Fuzhou, Fujian, China; Fujian Provincial Clinical Research Center for Cardiovascular Diseases Heart Center of Fujian Medical University, Fuzhou, Fujian, China; Department of Epidemiology and Health Statistics, School of Public Health, Fujian Medical University, Fuzhou, Fujian, China; Department of Medical Epidemiology and Biostatistics, Karolinska Institutet, Stockholm, Sweden

## Abstract

**Background:** Low-density lipoprotein cholesterol (LDLc) and body mass index (BMI) are not always correlated and their relationship is probably dependent on age, indicating differential age-specific effects of these two factors on health outcomes. We thus aimed to discriminate the effects of LDLc and BMI on the risk of coronary heart disease (CHD) across different age groups.

**Methods:** This is a prospective cohort study of 368,274 participants aged 38-73 years and free of CHD at baseline. LDLc and BMI were measured at baseline, and incident CHD was the main outcome. Cox proportional hazards model and restricted cubic spline (RCS) regression were used to estimate hazard ratio (HR) and 95% confidence interval (CI) of exposure on CHD.

**Results:** Similar relationships of LDLc and BMI with CHD risk were observed in overall population but in differential age-specific patterns. Across the age groups of <50, 50-54, 55-59, 60-64 and >=65 years, the LDLc-CHD association diminished with the adjusted HRs decreasing from 1.35, 1.26, 1.19, 1.11 to 1.08; while no declining trend was found in BMI-CHD relationship with the adjusted HRs of 1.15, 1.11, 1.12, 1.13 and 1.15, respectively. The interaction and mediation between LDLc and BMI on CHD risk were only present at young-age groups. And the LDLc-CHD but not BMI-CHD association was dependent on sex, metabolic syndrome and lipid-lowering drugs use.

**Conclusion:** There were differential age-specific effects of LDLc and BMI on the risk of developing CHD, calling for future efforts to discriminate the age-different benefits from lipids management or weight control on the primary prevention for CHD.

**Clinical Perspective:** 

**What Is New?:**

- This study shows, for the first time, that there were differential age-specific effects of LDLc and BMI on the risk of developing coronary heart disease (CHD), in which the LDLc-CHD association diminished with increasing age while no declining trend was found in BMI-CHD relationship.
- The interaction and mediation between LDLc and BMI on the risk of coronary heart disease were only present in young adults.

**What Are the Clinical Implications?:**

- Future efforts are called to discriminate the age-different benefits from lipids management or weight control on the primary prevention for CHD.
- Findings from this study highlighted that lipids management at young age was expected to have a much more favorable benefit in CHD prevention; and weight control shouldn’t be overlooked, especially for elderly adults, as lipid-lowering therapy might not contribute to the reduction of obesity-related CHD.

## Introduction

Population aging leads to an increased incidence of coronary heart disease (CHD). Low-density lipoprotein cholesterol (LDLc) and body mass index (BMI) are well-established determinants of CHD, both of which are complex phenotypes and shared lots of common risk factors. Besides, both obesity and high cholesterol have been involved in “paradox” phenomenon, defined as higher cholesterol or BMI being associated with an elevated incidence of diseases while paradoxically related to a reduced mortality ^1, 2^. Thus, it seems plausible to expect similar effects of these two factors on diseases. Nevertheless, emerging evidence found that LDLc and BMI were not always correlated, for example, findings from two large cross-sectional studies found that the association between BMI and LDLc was completely lost in population older than 60 years, indicating that the role of LDLc and BMI in diseases might not always stay in sync and probably varied by age ^3^. A number of studies have investigated the age-specific effect of LDLc on CHD but generated inconsistent results, and the influence of age was poorly demonstrated on the association between BMI and CHD ^4-9^. To date, there is insufficient data discriminating the independent effect of LDLc and BMI on the risk CHD across the lifespan.

The high prevalence of hyperlipidemia and obesity, as well as the demographic variation relevant to aging population, call for concerns to clarify the age-specific health effects of LDLc and BMI so as to anticipate benefits that might be achieved via LDLc reduction or weight control at different ages. Hence, in this study, we aimed to explore how the effects of LDLc and BMI on the risk of developing CHD varied from mid-life through late-life. Given of the prior evidence regarding the impact of sex and metabolic disorders in the BMI-LDLc relationship and the known effect of lipid-lowering treatment on CHD, we also performed stratified analyses to explore whether the age-specific associations differ by sex, metabolic syndrome (MetS) and lipid-lowering drug use ^3, 10^.

## Methods

### Study design and population

This cohort study was conducted based on UK Biobank project, which recruited more than 500 000 residents aged 38–73 years in 22 assessment centers across the UK ^11^. At the baseline visit between March 2006 and July 2010, participants completed online or/and verbal questionnaires and physical measurements, as well as provided biological sample. The North West Multi-Centre Research Ethics Committee has approved UK Biobank study (REC reference: 21/NW/0157) and all participants signed informed consent.

In this study, participants were excluded if they: (1) had a history of CHD before baseline visit, (2) had less than 180 days of follow-up, (3) had missing data of lipids or physical measurements. Finally, a total of 368,274 participants were included for analysis (**Supplementary Figure 1**).

### Assessment of exposure and covariates

Weight and height were measured at baseline and BMI was calculated as weight (kg) being divided by height (m) squared. Blood samples were collected at baseline and the levels of serum lipids were measured using the Beckman Coulter AU5800 Platform, in which LDLc was measured directly and analyzed via enzymatic selective protection method ^12, 13^. To reduce systematic error, standardized procedures were conducted for each sample following the protocol and Internal Quality Control (IQC) was performed for each assay, details of which have been described elsewhere ^12, 13^.

Sociodemographic and lifestyle information were obtained by self-reported questionnaires. Age was calculated as the difference between the date of birth and baseline assessment. Ethnicity was dichotomized as British or not. Education was recorded as six categories and unknown group. Socioeconomic status was measured by the Townsend deprivation index assigned at baseline ^14^. Smoking status was categorized as never, former smoker, current smoker and unknown status; passive smoking status was dichotomized as yes or not. Alcohol intake frequency was categorized as daily, often (one to four times a week), seldom (one to three times a month or special occasions only), never and unknown. Physical activity was assessed using the International Physical Activity Questionnaire and recorded as summed metabolic equivalent (MET) minutes per week for all activity ^15, 16^. Metabolic disorders, including hypertension, hyperglycemia/diabetes, hypertriglyceridemia and low high-density lipoprotein (HDL) cholesterol, were defined according to the IDF definition by combining information from self-report medical conditions, medications and biochemistry data at baseline (systolic blood pressure>=130 mmHg or diastolic blood pressure>=85 mmHg; fasting plasma glucose >=5.6 mmol/l; triglycerides>=1.7 mmol/l; HDL cholesterol<=1.03 mmol/l in men or <=1.29 mmol/l in women) ^17^. MetS was defined as having central obesity (waist circumference >=90 cm in men or >=80 cm in women) and two or more metabolic disorders mentioned above ^17^. Use of lipid-lowering drugs was self-reported in touchscreen questionnaire or verbal interview at baseline.

### Assessment of outcomes

The main outcome was incident CHD and the secondary outcome was incident myocardial infarction (MI). CHD was identified by using the International Classification of Disease 10 of I20-I25 from category of “First occurrence” (Category ID: 1712) with Field IDs of 131297, 131299, 131301, 131303, 131305 and 131307. Myocardial infarction was identified by using Field ID of 42001 from category of “Algorithmically-defined outcomes” (Category ID: 42). Fields in both “First occurrence” and “Algorithmically-defined outcomes” categories were derived from self-reported medical conditions, primary health care, hospital admission data and death registry data in the UK Biobank cohort.

### Statistical analysis

Baseline characteristics are presented as frequency and percentage for categorical variables and as mean and standard deviation for continuous variables across different age groups (i.e., <50 years, 50-54 years, 55-59 years, 60-64 years, and >=65 years). Participants were followed up from the date of baseline assessment to the date of the outcome occurrence, the date of death, the date of loss-to-follow, or the censoring date (May 20, 2022), whichever occurred first.

We first explored the associations between LDLc, BMI and risk of CHD in different age groups. Restricted cubic spline (RCS) regression was first applied to explore the non-linear relationship by including the exposure as a continuous variable, in which we tried to perform models with different knots and found that there was little change in the shape since four knots, thus RCS regression with four knots was chosen as the main model ^18^. Next, Cox proportional hazards model was used to investigate the hazard ratio (HR) and 95% confidence interval (CI) between LDLc, BMI and risk of CHD, in which, based on clinical relevance and previous literature, we modelled LDLc per 1 mmol/L increment as follows: <2, 2-2.9, 3-3.9, 4-4.9, 5-5.9, and ≥6 mmol/L; and modelled BMI per class increment using standard categories as follows: underweight (<18.5 kg/m^2^), healthy weight (18.5-24.9 kg/m^2^), overweight (25.0-29.9 kg/m^2^), obesity class 1 (30.0-34.9 kg/m^2^), obesity class 2 (35.0-39.9 kg/m^2^), and obesity class 3 (≥40 kg/m^2^) ^5, 19^. Given of the potential impact of competing risk from death, Fine-Gray competing risk model was further performed to examine the association by defining death as a competing event. Stratified analyses were also performed by sex, MetS or lipid-lowering drugs use across different age groups, and heterogeneity between strata was tested by including a product term of the stratification factor and exposure in the model. The models were first adjusted for age at baseline and sex, then in multivariable models additionally for ethnicity, education, smoking, passive smoking, alcohol intake frequency, physical activity, Townsend deprivation index, metabolic disorders, lipid lowering drugs, and body mass index or LDLc.

We further evaluated the age-specific mediation and interaction between LDLc and BMI on CHD. Mediation analysis was performed to explore the mediated role of LDLc between BMI and CHD, or the role of BMI between LDLc and CHD, by using the percentage of excess risk mediated (PERM), which was calculated by using the formula of (HR_(confounders adjusted)_ - HR_(confounders and mediator adjusted)_)/ (HR_(confounders adjusted)_ – 1). Interaction between LDLc and BMI on CHD were assessed in both additive scale and multiplicative scale ^20^. Addictive interaction was measured by relative excess risk due to interaction (RERI), calculated as HR_(increased LDLc & increased BMI)_ - HR_(increased BMI alone)_ – HR_(increased LDLc alone)_ + 1, in which LDLc and BMI was dichotomized as increased or not according to the cut-off value of 3.4 mmol/L and 25 kg/m^2^ respectively, and then the population was divided into four groups for analysis (i.e., neither LDLc nor BMI increased, LDLc increased alone, BMI increased alone and both increased). Multiplicative interaction was identified by the p value of product-term of LDLc and BMI in the multivariable model.

All analyses were performed in SAS 9.4 or R 4.1.2 and *P*-value of <0.05 was considered as of statistical significance.

## Results

As shown in **Table 1**, a total of 368,274 participants were included in the analyses, consisting of 202,853 (55.1%) females and 165,421 (44.9%) males. The mean age of study population was 57 years, the mean level of LDLc was 3.6 mmol/L and the mean BMI was 27.3 kg/m^2^. After a mean of 12 years of follow-up, 26,947 participants developed CHD with the incidence rate of 5.86 per 1000 person-years, and the incidence rates of CHD continuously increased with increasing age. There were 84717, 56544, 65680, 87263, 74070 participants across age groups of <50 years, 50-54 years, 55-59 years, 60-64 years, and >=65 years, and the distribution of sociodemographic characteristics, lifestyle, history of metabolic disorders and lipid-lowering drug use were significantly different across age groups. **Supplementary Figure 2** shows that participants with LDLc of 3-3.9 mmol/L or BMI of 25-29.5 kg/m^2^ accounted for the highest proportion across all age groups.

**Table 1.** Baseline information of the study population across age groups.

| **Variables** | **Study population**<br>**(N=368274)** | **<50 years**<br>**(N=84717)** | **50-54 years**<br>**(N=56544)** | **55-59 years**<br>**(N=65680)** | **60-64 years**<br>**(N=87263)** | **>=65 years**<br>**(N=74070)** | **P**<br>**value** |
| --- | --- | --- | --- | --- | --- | --- | --- |
| Age | 56.7±8.1 | 45.2±2.6 | 52.1±1.4 | 57.1±1.4 | 62±1.4 | 67.1±1.6 | <0.001 |
| LDL Cholesterol, mmol/L | 3.6±0.8 | 3.4±0.8 | 3.6±0.8 | 3.7±0.8 | 3.7±0.9 | 3.6±0.9 | <0.001 |
| Body mass index, kg/m<sup>2</sup> | 27.3±4.7 | 26.9±4.9 | 27.3±4.9 | 27.4±4.8 | 27.4±4.6 | 27.4±4.3 | <0.001 |
| Townsend deprivation index | -1.4±3 | -0.9±3.2 | -1.2±3.1 | -1.4±3 | -1.6±2.9 | -1.6±2.9 | <0.001 |
| Summed MET, min/week | 2668±2718 | 2660.7±2769.7 | 2569.4±2719.2 | 2538.7±2691.5 | 2706.4±2693.2 | 2829.1±2696.5 | <0.001 |
| Sex |  |  |  |  |  |  | <0.001 |
| Female | 202853(55.1) | 45922(54.2) | 31962(56.5) | 37048(56.4) | 48783(55.9) | 39138(52.8) |  |
| Male | 165421(44.9) | 38795(45.8) | 24582(43.5) | 28632(43.6) | 38480(44.1) | 34932(47.2) |  |
| Ethnicity |  |  |  |  |  |  | <0.001 |
| British | 324425(88.1) | 69723(82.3) | 48715(86.2) | 58283(88.7) | 79975(91.6) | 67729(91.4) |  |
| Others | 43849(11.9) | 14994(17.7) | 7829(13.8) | 7397(11.3) | 7288(8.4) | 6341(8.6) |  |
| Education |  |  |  |  |  |  | <0.001 |
| College or University degree | 121308(32.9) | 33372(39.4) | 20958(37.1) | 23554(35.9) | 25834(29.6) | 17590(23.7) |  |
| A levels/AS levels or equivalent | 41605(11.3) | 11586(13.7) | 7516(13.3) | 7785(11.9) | 8639(9.9) | 6079(8.2) |  |
| O levels/GCSEs or equivalent | 79500(21.6) | 19824(23.4) | 11953(21.1) | 13166(20) | 18968(21.7) | 15589(21) |  |
| CSEs or equivalent | 20785(5.6) | 8704(10.3) | 4737(8.4) | 3660(5.6) | 2453(2.8) | 1231(1.7) |  |
| NVQ or equivalent | 23660(6.4) | 4151(4.9) | 3627(6.4) | 4642(7.1) | 6419(7.4) | 4821(6.5) |  |
| Other professional qualifications | 18785(5.1) | 2118(2.5) | 2107(3.7) | 3430(5.2) | 5681(6.5) | 5449(7.4) |  |
| Unknown | 62631(17) | 4962(5.9) | 5646(10) | 9443(14.4) | 19269(22.1) | 23311(31.5) |  |
| Smoking |  |  |  |  |  |  | <0.001 |
| Never | 203911(55.4) | 51766(61.1) | 33515(59.3) | 36140(55) | 45370(52) | 37120(50.1) |  |
| Previous | 124601(33.8) | 20813(24.6) | 16121(28.5) | 22578(34.4) | 33955(38.9) | 31134(42) |  |
| Current | 37998(10.3) | 11785(13.9) | 6702(11.9) | 6688(10.2) | 7497(8.6) | 5326(7.2) |  |
| Unknown | 1764(0.5) | 353(0.4) | 206(0.4) | 274(0.4) | 441(0.5) | 490(0.7) |  |
| Passive smoking |  |  |  |  |  |  | <0.001 |
| No | 322151(87.5) | 71988(85) | 48725(86.2) | 57536(87.6) | 77708(89.1) | 66194(89.4) |  |
| Yes | 46123(12.5) | 12729(15) | 7819(13.8) | 8144(12.4) | 9555(10.9) | 7876(10.6) |  |
| Alcohol intake frequency |  |  |  |  |  |  | <0.001 |
| Daily | 75829(20.6) | 12803(15.1) | 10445(18.5) | 14162(21.6) | 20596(23.6) | 17823(24.1) |  |
| Often | 180605(49) | 44256(52.2) | 29248(51.7) | 32753(49.9) | 41565(47.6) | 32783(44.3) |  |
| Seldom | 82719(22.5) | 20912(24.7) | 12682(22.4) | 13973(21.3) | 18454(21.1) | 16698(22.5) |  |
| Never | 28341(7.7) | 6510(7.7) | 4041(7.1) | 4667(7.1) | 6511(7.5) | 6612(8.9) |  |
| Unknown | 780(0.2) | 236(0.3) | 128(0.2) | 125(0.2) | 137(0.2) | 154(0.2) |  |
| Metabolic disorder |  |  |  |  |  |  |  |
| Hypertension | 237905(64.6) | 38136(10.4) | 32477(8.8) | 42877(65.3) | 64124(73.5) | 60291(81.4) | <0.001 |
| Hyperglycemia/diabetes | 59777(16.2) | 9170(2.5) | 7578(2.1) | 10562(16.1) | 16207(18.6) | 16260(22) | <0.001 |
| Hypertriglyceridemia | 145646(39.5) | 28048(7.6) | 21298(5.8) | 26693(40.6) | 37539(43) | 32068(43.3) | <0.001 |
| Low HDL Cholesterol | 74743(20.3) | 20459(5.6) | 11467(3.1) | 12254(18.7) | 16380(18.8) | 14183(19.1) | <0.001 |
| Use of lipid lowering drugs | 51240(13.9) | 3082(0.8) | 4360(1.2) | 8032(12.2) | 16337(18.7) | 19429(26.2) | <0.001 |

As shown in **Figure 1 and Supplementary Figure 3**, RCS regression found near linear relationship between LDLc, BMI and CHD risk, especially when the level of LDLc or BMI was above 3 mmol/L or 25 kg/m^2^. Stratified analyses by age groups observed that the association of LDLc but not BMI with CHD almost diminished with the increasing age (**Figure 2**). Cox proportional hazards model found similar elevated risk of CHD for the overall population per 1 mmol/L increment in LDLc (multivariable adjusted HR: 1.16, 95% CI: 1.14-1.18) and per class increment in BMI (multivariable adjusted HR: 1.14, 95% CI: 1.12-1.16) (**Table 2**). Consistent with findings from RCS regression, stratified analyses in Cox proportional hazards model showed that across the age groups of <50, 50-54, 55-59, 60-64 and >=65 years, the positive association between LDLc and CHD declined along the increasing age with the multivariable adjusted HRs decreasing from 1.35, 1.26, 1.19, 1.11 to 1.08 (*P*_heterogeneity_<0.001); however, although there was a significant heterogeneity in BMI-CHD associations across age groups, no decling trend was observed with increasing age, and the multivariable adjusted HRs of which were 1.15, 1.11, 1.12, 1.13 and 1.15, respectively (**Figure 3 and Table 2**). Fine-Gray competing risk model showed similar results, suggesting a small role of the competing risk in the observed associations (**Supplementary Table 1**). **Supplementary Table 2** shows the results for MI; similarly, the positive association of LDLc with the risk of MI also declined across the increasing age (*P*_heterogeneity_<0.001) while the relationship between BMI and MI was independent of age (*P*_heterogeneity_=0.128). Although the effect on MI related to per 1 mmol/L higher in LDLc was always greater than that associated with per class higher in BMI across all age groups, the difference in the association of LDLc and BMI with MI was attenuating with increasing age.

**Figure 1.**
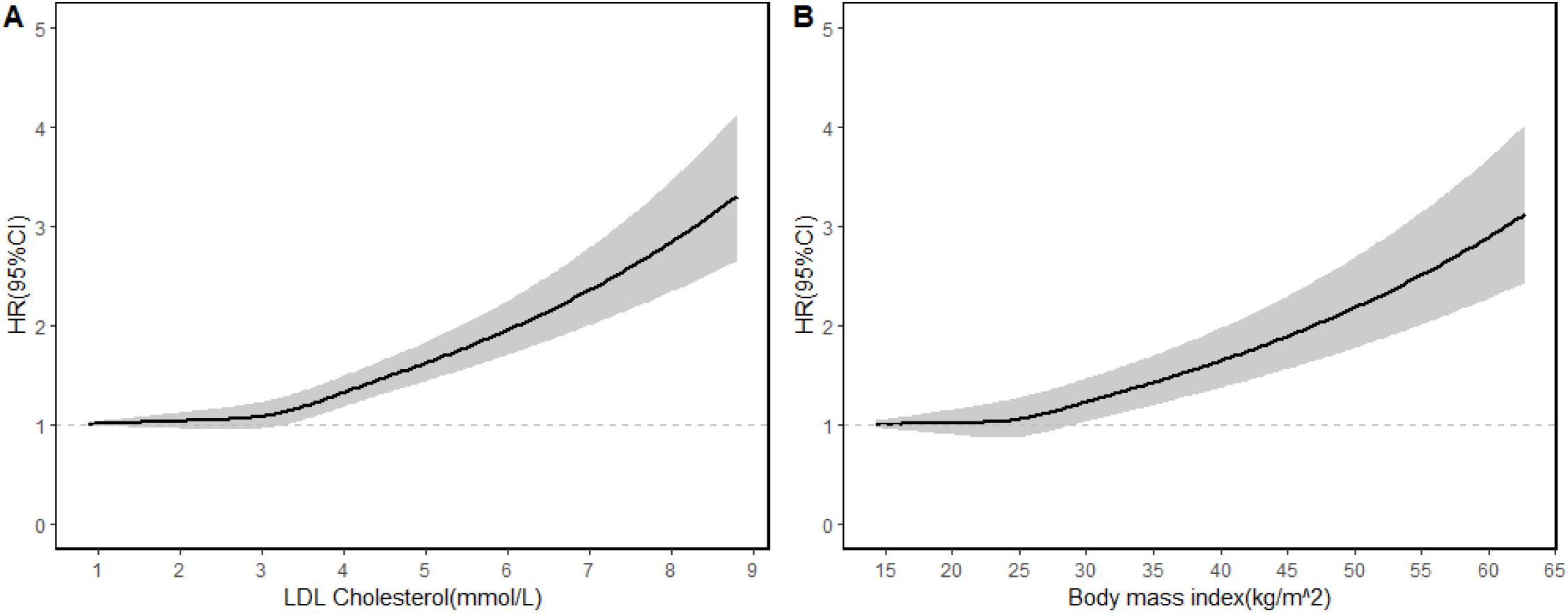
Association of LDL cholesterol or body mass index with risk of coronary heart disease via restricted cubic spline regression (four knots). *Adjusted for age, sex, ethnicity, education, smoking, passive smoking, alcohol intake frequency, physical activity, Townsend deprivation index, metabolic disorders, lipid lowering drugs and body mass index or LDL cholesterol.

**Figure 2.**
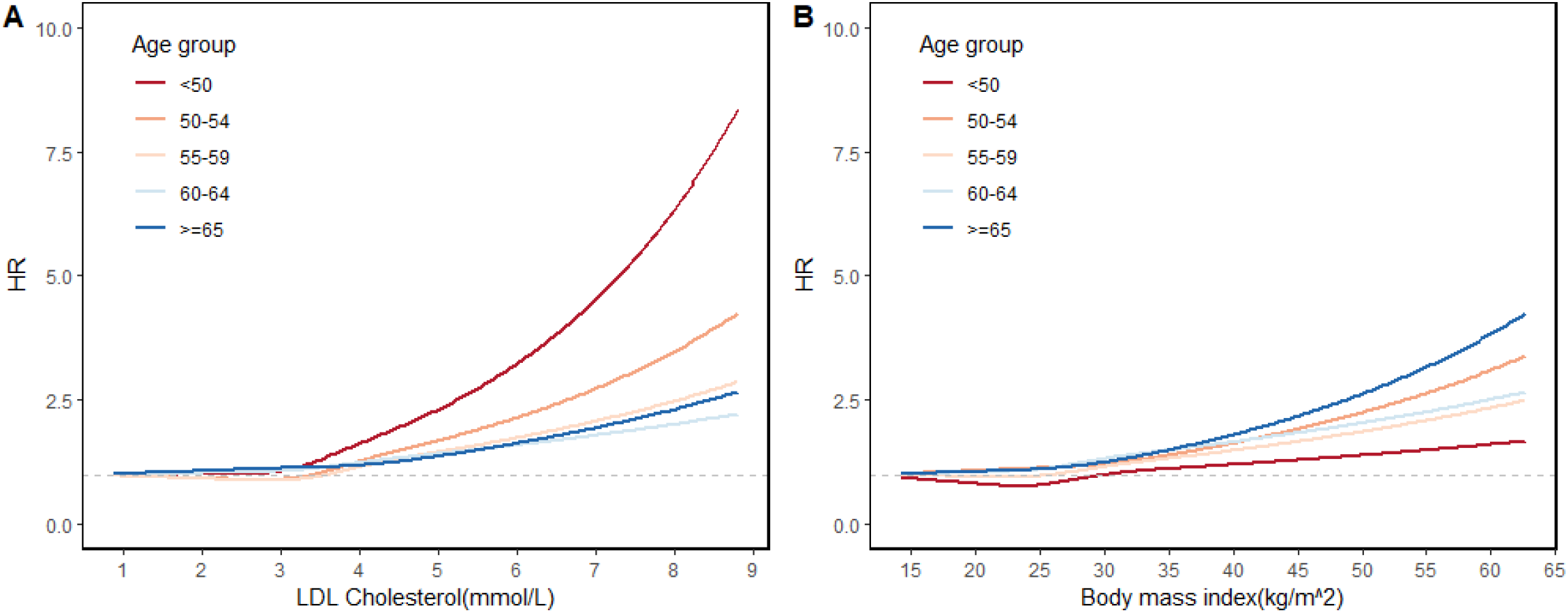
Association of LDL cholesterol or body mass index with risk of coronary heart disease across age groups via restricted cubic spline regression (four knots). *Adjusted for age, sex, ethnicity, education, smoking, passive smoking, alcohol intake frequency, physical activity, Townsend deprivation index, metabolic disorders, lipid lowering drugs and body mass index or LDL cholesterol.

**Figure 3:**
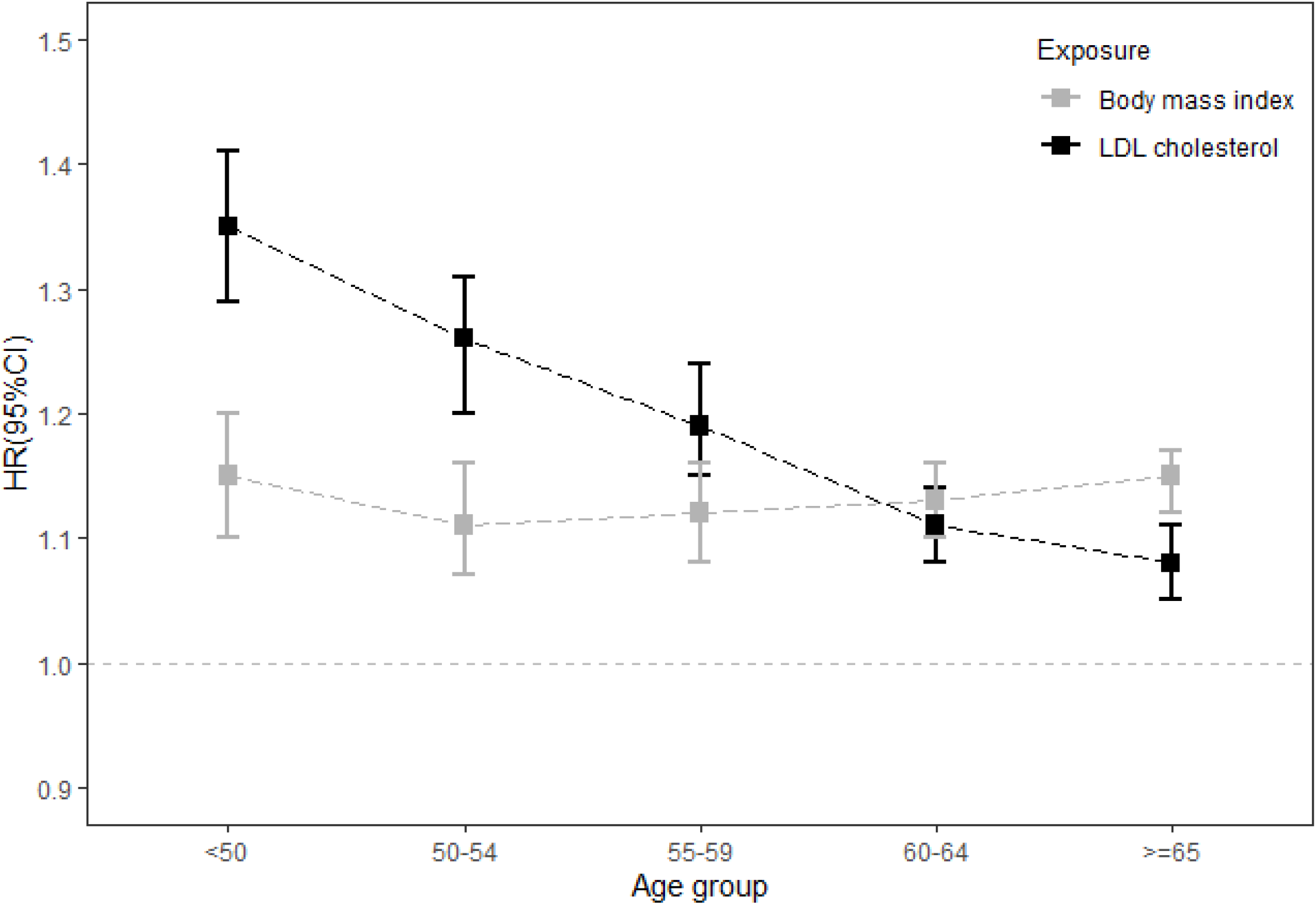
Association of LDL cholesterol or body mass index with risk of coronary heart disease across increasing age via Cox proportional hazards model. *Adjusted for age, sex, ethnicity, education, smoking, passive smoking, alcohol intake frequency, physical activity, Townsend deprivation index, metabolic disorders, lipid lowering drugs and body mass index or LDL cholesterol.

**Table 2.** Association of LDL cholesterol or body mass index with risk of coronary heart disease across age groups.

| Age | Number of individuals | Number of outcomes | Follow up years | IR, per 1000 person years | LDL cholesterol |  | Body mass index |  |
| --- | --- | --- | --- | --- | --- | --- | --- | --- |
|  |  |  |  |  | Age and sex adjusted | Multivariable adjusted<sup>a</sup> | Age and sex adjusted | Multivariable adjusted<sup>a</sup> |
| Overall | 368274 | 26947 | 4595322 | 5.86 | 1.04(1.02,1.05) | 1.16(1.14,1.18) | 1.31(1.29,1.32) | 1.14(1.12,1.16) |
| <50 | 84717 | 2524 | 1099075 | 2.30 | 1.38(1.32,1.44) | 1.35(1.29,1.41) | 1.46(1.40,1.51) | 1.15(1.10,1.20) |
| 50-54 | 56544 | 2746 | 723131 | 3.80 | 1.18(1.13,1.23) | 1.26(1.20,1.31) | 1.33(1.28,1.38) | 1.11(1.07,1.16) |
| 55-59 | 65680 | 4361 | 827492 | 5.27 | 1.06(1.03,1.10) | 1.19(1.15,1.24) | 1.31(1.28,1.35) | 1.12(1.08,1.16) |
| 60-64 | 87263 | 7928 | 1071790 | 7.40 | 0.99(0.96,1.01) | 1.11(1.08,1.14) | 1.28(1.25,1.31) | 1.13(1.10,1.16) |
| >=65 | 74070 | 9388 | 873834 | 10.74 | 0.94(0.92,0.96) | 1.08(1.05,1.11) | 1.27(1.24,1.30) | 1.15(1.12,1.17) |
| P heterogeneity |  |  |  |  | <0.001 | <0.001 | <0.001 | <0.001 |

<sup>a</sup> Adjusted for age, sex, birth country, education, smoking, passive smoking, alcohol intake frequency, physical activity, Townsend deprivation index, metabolic disorders, lipid lowering drugs and body mass index or LDL cholesterol.

In **Figure 4 and Supplementary Table 3**, stratified analyses by sex found a significantly stronger LDLc-CHD association (P heterogeneity for overall population: <0.001) and a slightly insignificantly stronger BMI-CHD association (P heterogeneity for overall population: 0.405) in males than females across all age groups; irrespective of sex, the LDLc-CHD association declined with increasing age while the BMI-CHD association kept stable. In **Figure 2 and Supplementary Table 4**, stratified analyses by MetS observed a stronger association between LDLc and CHD among individuals without MetS than those with, across all age groups despite without statistical significance in some age groups (P heterogeneity for overall population: <0.001), while no significant heterogeneity was found in BMI-CHD association between individuals with and without MetS in most age groups except the age group of younger than 50 years (P heterogeneity for overall population: 0.346); regardless of MetS, diminished trend was found in LDLc-CHD but not BMI-CHD association with increasing age. In **Figure 2 and Supplementary Table 5**, stratified analyses by lipid-lowering drug use showed significant heterogeneity in LDLc-CHD association between lipid-lowering drug users and non-users, specifically among adults younger than 55 years old (P heterogeneity=0.001); besides, the LDLc-CHD association declined with increasing age in non-users but kept stable in lipid-lowering drug users, while no obvious trend was observed for BMI-CHD association regardless of lipid-lowering drug use or not.

**Figure 4:**
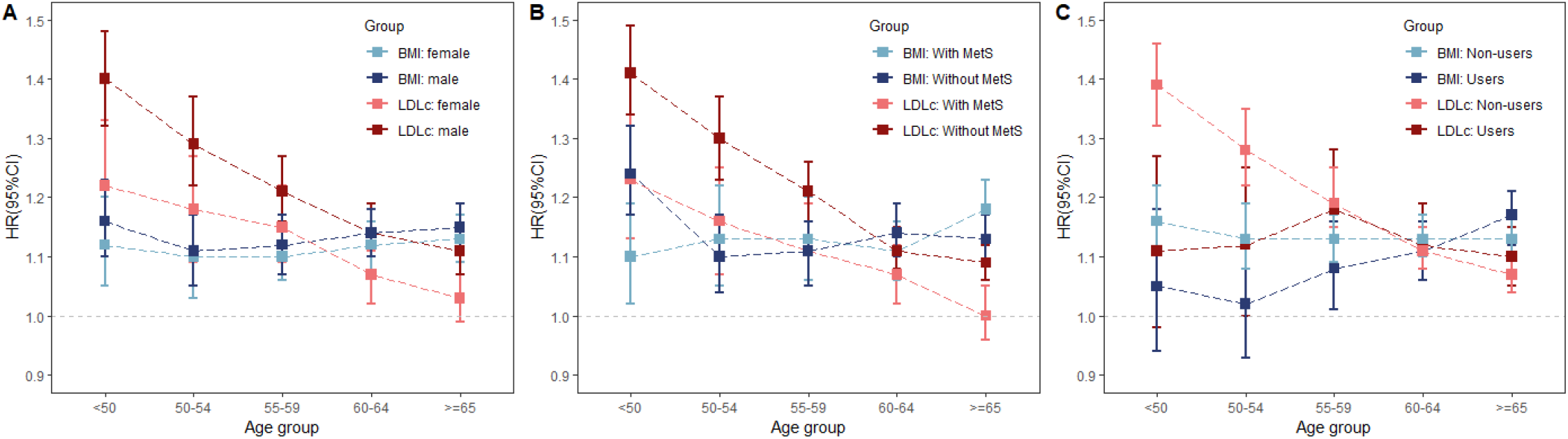
Stratified analyses for the association of LDL cholesterol or body mass index with risk of coronary heart disease across age groups via Cox proportional hazards model. *Adjusted for age, sex, ethnicity, education, smoking, passive smoking, alcohol intake frequency, physical activity, Townsend deprivation index, metabolic disorders, lipid lowering drugs and body mass index or LDL cholesterol.

Mediation analysis found that LDLc played a mediated role in BMI-CHD association at young ages with the PERM of 11.76% and 8.33% in the age group of <50 years and 50-54 years respectively; the mediated effect of BMI in LDLc-CHD association was only observed in the age group of <50 years with the PERM of 2.78% (**Table 3)**. Interaction analysis showed a greater positive addictive interaction between BMI and LDLc in younger adults than older ones although no significant multiplicative interaction found throughout all ages; the effect of higher LDLc alone or both higher BMI and LDLc on CHD risk declined with increasing age while no significant trend was found for the effect of higher BMI alone on CHD (**Table 4)**.

**Table 3.** Mediation of LDL cholesterol or body mass index between body mass index or LDL cholesterol and risk of coronary heart disease across age groups.

| Age | LDL cholesterol |  |  | Body mass index |  |  |
| --- | --- | --- | --- | --- | --- | --- |
|  | Confounders adjusted<sup>a</sup> | Confounders and BMI adjusted<sup>b</sup> | The percentage of excess risk mediated | Confounders adjusted<sup>a</sup> | Confounders and LDLc adjusted<sup>b</sup> | The percentage of excess risk mediated |
| Overall | 1.16(1.15,1.18) | 1.16(1.14,1.18) | 0% | 1.14(1.13,1.16) | 1.14(1.12,1.16) | 0% |
| <50 | 1.36(1.30,1.42) | 1.35(1.29,1.41) | 2.78% | 1.17(1.12,1.21) | 1.15(1.10,1.20) | 11.76% |
| 50-54 | 1.26(1.20,1.32) | 1.26(1.20,1.31) | 0% | 1.12(1.08,1.17) | 1.11(1.07,1.16) | 8.33% |
| 55-59 | 1.19(1.15,1.24) | 1.19(1.15,1.24) | 0% | 1.12(1.09,1.16) | 1.12(1.08,1.16) | 0% |
| 60-64 | 1.11(1.08,1.14) | 1.11(1.08,1.14) | 0% | 1.13(1.10,1.16) | 1.13(1.10,1.16) | 0% |
| ≥65 | 1.07(1.05,1.10) | 1.08(1.05,1.11) | - | 1.15(1.12,1.17) | 1.15(1.12,1.17) | 0% |

<sup>a</sup> Adjusted for age, sex, ethnicity, education, smoking, passive smoking, alcohol intake frequency, physical activity, Townsend deprivation index, metabolic disorders, lipid lowering drugs;
<sup>b</sup> Adjusted for age, sex, ethnicity, education, smoking, passive smoking, alcohol intake frequency, physical activity, Townsend deprivation index, metabolic disorders, lipid lowering drugs and body mass index or LDL cholesterol.

**Table 4.**
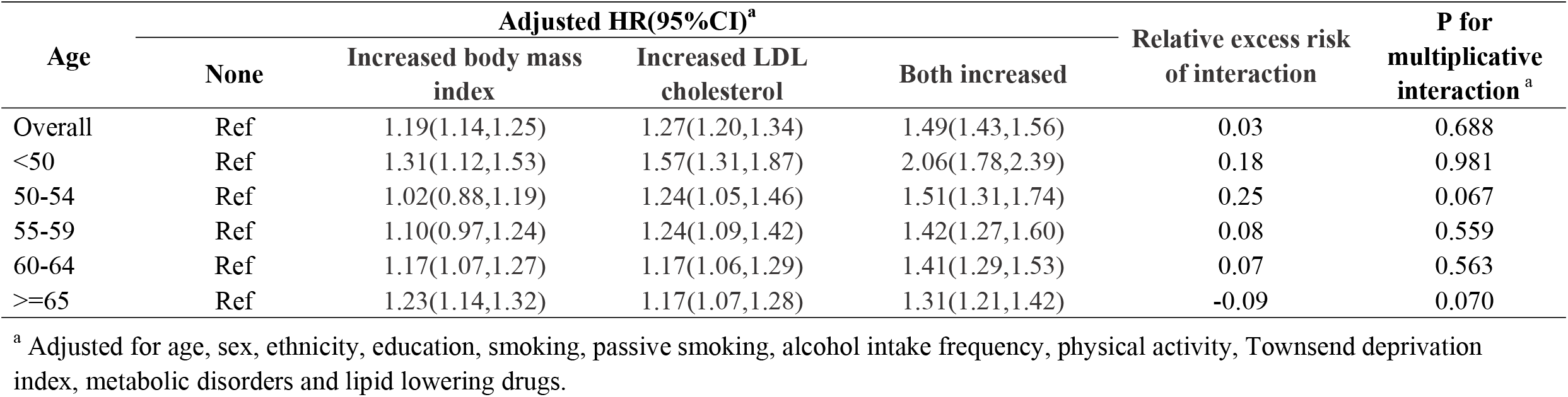
Interaction between LDL cholesterol and body mass index on the risk of coronary heart disease across age groups.

## Discussion

This prospective cohort study found similar independent effects of LDLc and BMI on CHD risk in overall population, while the age-specific associations with CHD risk showed significant different patterns between LDLc and BMI. The LDLc-CHD association declined with increasing age while the BMI-CHD relationship was independent of age. The strongest negative effect of high LDLc on CHD was found in young adults without using lipid-lowering drugs, and the interactive effect between LDLc and lipid-lowering drug use on CHD risk was only observed in adults younger than 55 years. Besides, the interaction and mediation between LDLc and BMI on CHD were only present in young adults. These findings highlighted that lipids management at young age was expected to have a much more favorable benefit in CHD prevention; and weight control shouldn’t be overlooked, especially for elderly adults, as lipid-lowering therapy might not contribute to the reduction of obesity-related CHD.

Hypercholesterolemia and obesity share some common risk factors and pathophysiologic mechanisms, both of non-negligible importance in the onset of CHD. Although previous literature has found observational and genetic associations between BMI and LDLc, growing evidence suggested that the correlation seemed to be varied by metabolic status linked to aging or metabolic disorders, indicating that the health effect of BMI and LDLc might not always stay consistent^3, 10, 21, 22^.

Age has been reported to play a role in the association of BMI or LDLc with health outcomes ^1, 4-8^. Despite the causal relationship between LDLc and the risk of CHD, the impact of LDLc or the benefit of lipid-lowering therapy on the occurrence of CHD is being challenged in older population^4-8, 23-25^. Some studies have observed that the positive association of LDLc with incident CHD attenuated or even disappeared in older adults, while a recent contemporary cohort argued that individuals aged 70-100 years with higher LDLc had the highest absolute risk of atherosclerotic cardiovascular disease as compared to other age groups, suggesting the importance of LDLc-lowering treatment for prevention in extremely-elderly population^4-8^. The net benefit of statins use on primary prevention of atherosclerotic cardiovascular events was also controversial and guidelines regarding the role of statins remained equivocal in older adults^23-25^. The “obesity paradox” phenomenon has been proposed for decades with respect to the lower mortality in obese adults in older patients ^1^. Nevertheless, the age-related effect of obesity in the incidence of diseases was rarely clarified, and previous studies found a declining trend in the effect of higher BMI on the occurrence of some diseases, such as dementia, while a cohort study of population aged >60 years found that obesity was independently associated with elevated incidence of CHD in older population, at least to age 84 ^9, 26^. There is a dearth of data on discrimination of the independent effect of LDLc and BMI on health outcomes across age groups. In the current study, we found similar relationships for both LDLc and BMI with the risk of developing CHD but differential age-specific effects between LDLc and BMI. The excess risk on CHD from higher LDLc attenuated with increasing age while the relationship between BMI and the incidence of CHD kept stable across all age groups, resulting in a greater excess risk associated with per mmol/L higher in LDLc in young adults but a higher excess risk related to per class increase in BMI in adults older than 60 years. Despite declined strength of the LDLc-CHD association in elderly people, the negative health effect of higher LDLc stayed statistically significant. These findings casted doubt on suggestions about guideline revision given of the “obesity paradox” or “cholesterol paradox” phenomenon at older ages but called for future efforts to discriminate the age-different benefits from lipids management or weight control on the primary prevention for CHD.

Some studies have investigated the mediated role of LDLc between BMI and CHD, for example, a large pooled analysis found the excess risk of CHD per unit of BMI can be largely explained by metabolic mediators whereas cholesterol explained only additional 5% excess risk; another population-based study found that LDLc explained the excess risk of CHD with 22% excess risk from the observational obesity and 8% excess risk from genetically-determined obesity^22, 27^. In this study, we extended the knowledge in light of the age-different mediation or interaction between LDLc and BMI on the risk of CHD, and found that the mediated effect of LDLc or BMI between BMI or LDLc and CHD risk was only present in young adults, so did the interaction effect, indicating a greater benefit of lipids management or weight control on preventing CHD at young ages.

Difference in the sex- or MetS-specific effects was further observed between LDLc and BMI on CHD risk in this study. A prior cohort study in Korean population demonstrated sex-specific association of MetS (an obesity-driven complex disorder) and LDLc with the incidence of cardiovascular and cerebrovascular disease (CCVD), in which MetS alone was linked with a higher risk of CCVD regardless of sex while the impact of higher LDLc alone on CCVD was more pronounced in men than in women ^28^. Consistently, the current study found that the association of BMI with CHD risk was independent of sex but the effect of high LDLc on CHD was significantly greater in men than in women across all age groups. MetS also seemed to play a role in the LDLc-CHD but not BMI-CHD relationship; the effect of higher LDLc was stronger among people without MetS than those with, which called for more studies in light of the underlying reasons. In addition, stratified analyses by lipid-lowering drug use observed the highest excess risk of CHD related to high LDLc in youngest adults without using lipid-lowering drugs and the absent heterogeneity in the effect of LDLc on CHD between users and non-users in older adults, further suggesting that lipids management should start at earlier ages for the primary prevention of CHD.

This study had some strengths. First, this study extended the knowledge regarding the differential age-specific effects of LDLc and BMI on the risk of developing CHD. Second, the large sample size allowed us to explore the associations across age groups per 5-year interval with sufficient statistical power. Third, information of the outcome was obtained by combining several sources to minimize the misclassification of outcome. Fourth, several statistical methods were applied to estimate the associations, contributing to examine the robustness of the results. The major limitation of this study was that the age of this study population ranged from 38 to 73, thus not allowing us to explore the relationship in extreme elderly individuals. Besides, most participants in UK Biobank are European, thus findings should be interpreted cautiously in terms of generalization.

In conclusion, this study found differential age-specific effects between LDLc and BMI on the risk of developing CHD, calling for future efforts to discriminate the age-different benefits from lipids management or weight control on the primary prevention for CHD.

## Data Availability

Data from the UK Biobank is available to researchers upon a data access application.

## Abbreviations

BMI: body mass index
CHD: coronary heart disease
LDLc: low- density lipoprotein cholesterol
MetS: metabolic syndrome
MI: myocardial infarction
PERM: percentage of excess risk mediated
RCS: restricted cubic spline
RERI: relative excess risk due to interaction.

## Acknowledgments

This study has been performed using the UK Biobank Resource under Application Number 84352. We thank the subjects and investigators who contributed to the UK Biobank.

